# Post marketing surveillance for Microwave Treatment of Plantar and Common Warts in Adults

**DOI:** 10.1101/2022.02.08.22270290

**Authors:** Ivan Bristow, Shailesh Joshi, Jonathan Williamson, Michael Ardern-Jones

## Abstract

A handheld microwave device (Swift^®^, Emblation Limited) has been licenced and available for clinical use since 2016 in the fields of podiatry and dermatology and has been extensively used in treating cutaneous warts. As part of post marketing surveillance by the manufacturer, an online 79-item survey was distributed to podiatry clinics in the United Kingdom with a Swift^®^ device. A total of 126 clinics responded (59.6%). 6998 adults (<65 years) underwent wart treatment with microwave (81.9% plantar warts; 18.1% common, non-plantar warts). The median efficacy rate was reported as 79.2% (65.9 - 87.5%) and 82.3% (71.4 - 100%) respectively. In older adults (over 65 years) efficacy rates were similar for both sites: plantar (73.2%, 50-90%, n=1232) and non-plantar (80.0%, 42.1-100%, n=276). A median of three treatments was required to bring about resolution. Sub-group analysis of the data revealed good clearance rates in patients with diabetes (79.6%), but less in immunocompromised individuals (61.3%) and those with autoimmune disease (58.6%). Overall, mean user satisfaction was rated as “very satisfied” on a 10-point scale (n=93 practices). A small number of adverse events were reported including blistering, superficial ulceration and poor healing were reported (n=7). Despite the limitations of a post-marketing questionnaire survey, these data provide good evidence of the safety and efficacy for Swift^®^ microwave treatment of cutaneous warts.

**What is already known about this topic?:**

- Microwave treatment has shown to be effective in the treatment of cutaneous warts.

**What does the study add?:**

- This survey of clinics using SWIFT microwave treatments reports on the effectiveness and safety of the device in the treatment of 8506 adult patients with common and plantar warts.
- Responding user clinics reporting efficacy with good clearance rates and low levels of adverse events demonstrating microwave is a safe and effective treatment for plantar and common warts.

## Introduction

Cutaneous warts are a common infection of the epidermis caused by the Human Papilloma Virus (HPV). Despite a wide range of treatment being advocated, a meta-analysis of treatment outcomes has shown disappointing clearance rates when compared with placebo ^1^. In recent years there have been a number of papers exploring the use of heat as a treatment modality for HPV skin infection ^2,3^. Warts heated to around 41° – 44° Celsius have shown increased clearance versus untreated lesions ^4^. Microwave energy is a modality capable of rapidly increasing tissue temperatures through dielectric heating and is available for handheld skin directed use (Swift^®^ Emblation Limited, Alloa, Scotland). In preclinical work on human skin we have demonstrated that microwave therapy induces up regulation of Heat Shock Protein 70 (HSP70) and enhances dendritic cell presentation of HPV antigens to CD8+ T cells by keratinocytes. This results in increased T cell activation ^5^. A subsequent clinical study demonstrated a 75.9% clearance rate of recalcitrant plantar lesions in adults after four monthly applications of microwave energy to the lesions using the device ^6^. The device was launched to the general market in 2016 and here are the latest results from post marketing surveillance as required under the by EU Medical Devices regulations.

## Methods

Swift microwave users were identified from the manufacturer’s customer database and invited by email to complete a survey. An inducement to responders was offered to complete the survey of a future discount on servicing the swift unit was offered. The approximate time required to complete the survey was 45-60 minutes. There was a time limit of two months to complete the survey. Replies were captured using secure online survey software. Of the 79 questions (supplementary file 1), 52 were independent and 27 were dependent. 49 (62%) examined clinical use, 18 (23%) efficacy, and 7 (9%) patient satisfaction. Data was evaluated using Minitab^®^ 14.0 statistical software.

## Results

Of the 193 invitations sent, 126 responses (65.3%) were received to the online system (table 1). Of these 22 (17.4%) were duplicate responses received from 11 practices, equalling a questionnaire response rate of 59.6% of invited podiatry practices demonstrating a good range of experience with Swift by responders (table 2). The precise reason for duplicate responses was not discernible retrospectively but from the time of data entry recorded it appeared that in most cases, the second response provided more data suggesting that the first data entry had been halted to gather more data. Whilst, as a stringent approach to data analysis we excluded any practices with duplicate entries, sensitivity analysis showed that by including one of the duplicate entries (with the most data), the findings reported were not significantly altered. 11 responses (8.7%) were excluded because at least one of the responses supplied was incompatible with another response suggesting possible misunderstanding of the questions posed. Overall, data was entered for 77.5% (σ =0.16) of the independent questions (figure 1).

**Table 1.**
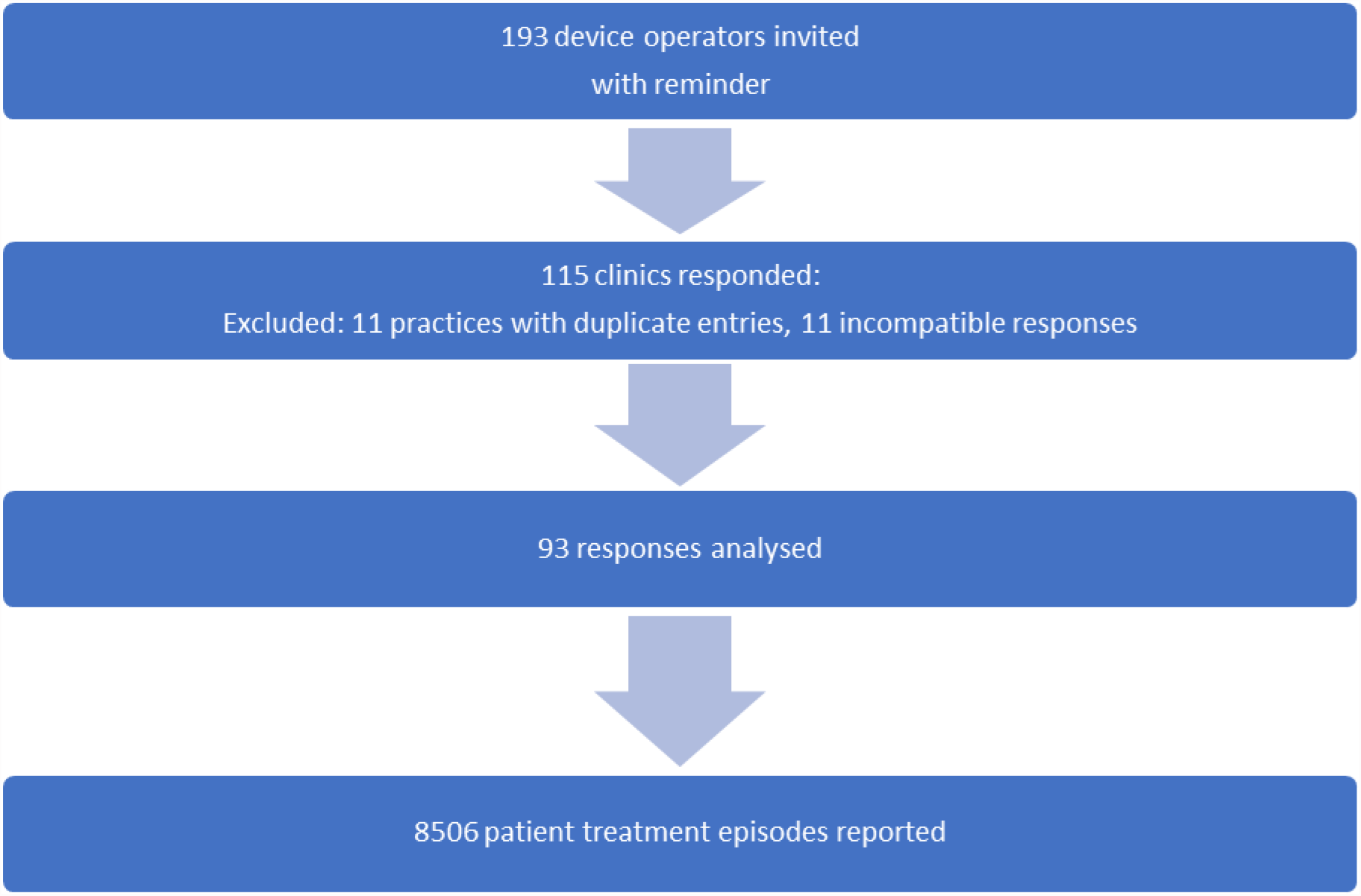
Summary of Responses.

**Table 2.** Experience with SWIFT reported by questionnaire responders.

| **Device use** | **% of Clinicians** |
| --- | --- |
| 3 years or more | 23% |
| Less than 3 years | 30% |
| Less than 2 years | 23% |
| 6 months-1 year | 18% |
| Less than 6 months | 6% |

## Reported Efficacy

In adults (less 65 years of age), 5733 (median 53.5 per responding practice) plantar wart patients and 1265 (median 6 per responding practice) common (non-plantar) wart patients had been treated with Swift. Analysis of efficacy from the retrospective questionnaire data was not possible on a per wart basis, therefore results are presented on a per patient basis. The median efficacy rate (defined as the number of resolved cases divided by the number of completed treatments) was reported as 79.2% (65.9 - 87.5%) and 82.3% (71.4 - 100%) respectively. Adults over 65 years of age had fewer patients treated (1232 plantar and 276 common), but showed similar resolution rates of 73.2% (50-90%) for plantar warts and 80.0% (42.1-100%) for common warts respectively (Table 3).

**Table 3.**
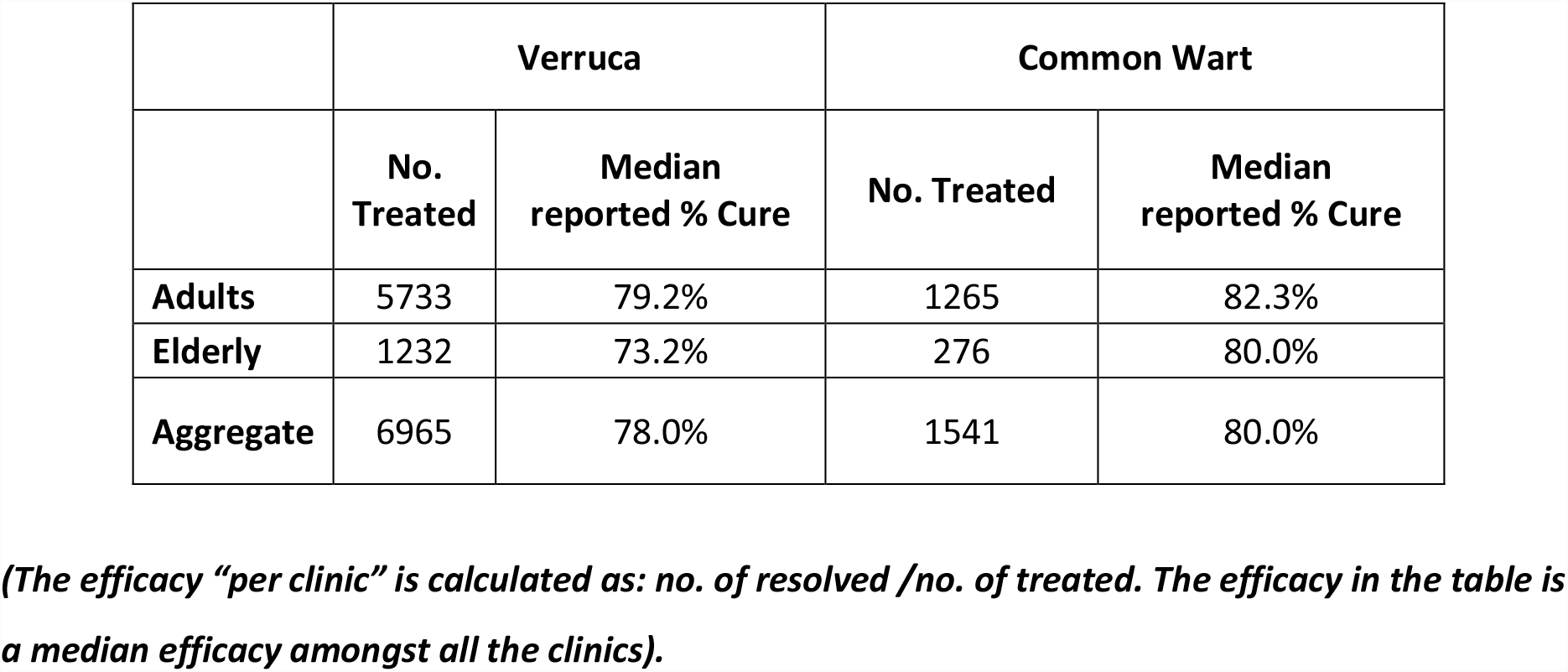
Reported Efficacy of SWIFT treatment.

81.8% (63 of 77) of the respondents did not report any instance of recurrence at the previously microwave treated sites. The remaining clinicians who observed any evidence of recurrence reported very low % of patients in comparison to the overall total number of patients treated. This figure (% of patients) is not included as it is not certain that all the patients were routinely contacted regarding recurrence.

## Treatment Regimes Employed

80.5% of respondents confirmed that they used the published treatment regime with 8-10 Watt energy dose delivered for 2 seconds (16-20 Joules), repeated 3 - 5 times at each appointment. Interestingly, 78.7% of clinicians reported adopting a modified treatment regimen for specific cases which typically reduced the energy per treatment (median 12 Joules, IQR 10-20 Joules) but increased the number of applications per site (median 6 repetitions, IQR 5-9.5). The reasons given for a varied treatment regime included circumstances such as long-standing lesions, aggressive lesions, pain threshold, different anatomical locations etc.

Respondents reported a median of 3 treatments was required to bring about resolution in responding lesions with treatment intervals of median 4 weeks and a median review period of 3 months. Of the respondents (84%) who described clinical data on treated lesions, 69.2% (n=54) reported treating all lesions present whilst 30.8% (n=24) reported treating only the primary lesion(s) leaving satellite lesions untreated. There was no evidence that efficacy was enhanced by treating all lesions at once (P > 0.05 for all analyses of those 18-65 years, or over 65 years with plantar and common warts, Mann-Whitney test). Of the 24 respondents treating only primary lesions, 16 (66.6%) reported that they had identified untreated lesions resolve without direct microwave treatment.

## Special patient groups

Respondents were requested to identify any patients with known conditions which may affect their treatment outcomes – patients with conditions known to compromise their immunity (such as drug therapies), patients with diabetes and those with auto-immune disorders. Within the subset, 52.1% reported having treated patients with immune-compromise, 45.3% diabetes and 30.7% autoimmune disease. Unsurprisingly, the reported response rates were lower in immunocompromised individuals (cohort clearance 61.3%, n=163) and those with autoimmune disease (cohort clearance 58.6%, n=75) but appear to be similar in those case with diabetes (cohort clearance 79.6%, n=206).

## Pain and adverse effects

Application of microwave energy is known to be painful for some patients. Clinicians were asked to document what methods they employed for reducing discomfort during treatment, if any. Within the group, 66.2% (n=47) clinicians actively sought to manage pain during the treatment. Approaches included deep breathing/mindfulness (37.6%), local anaesthetic (11.8%), topical anaesthetic (8.6%), Entonox (6.5%), gating or distraction techniques (4.3%) and ice (2.2%). Post-operatively, 37% (n=46) recommended oral analgesics such as paracetamol (96.3%) and less commonly ibuprofen (11.1%). 4/74 (5.4%) respondents reported that they had identified an occurrence of post-operative pain lasting over four weeks. 13 respondents (14.0%) entered data in the question recording experience of serious adverse effects. The descriptions were limited but 7 identified cases of poor healing, ulceration or blistering at the treatment site. One clinician identified two cases of lymphangitis which upon investigation (personal communication) were deemed non-device related.

## Satisfaction

Utilising a 10-point scale (0 very dissatisfied – 10 extremely satisfied), respondents were on average very satisfied (average 7.5; σ=1.87; n=93). Whilst indirectly reported via the practitioners, patient satisfaction was reported to be similar (7.31; σ=1.7; n=93). Respondents reported that they would recommend microwave treatment to 88.2% (σ=20.7%; n=93) of patients with a plantar wart.

## Discussion

This survey represents a review of clinical outcome data from 8506 treated common and plantar wart patients over a three-year period. The overall efficacy data for verrucae of 78.0% compares favourably with the resolution rate reported in the first clinical study of the device suggesting a clearance of 75.9% of lesions after 4 treatments or less in plantar warts ^6^.

Clearance of non-plantar lesions (common warts) has been shown to be similar to verrucae clearance rates or superior, using microwave (82.3% in adults under 65, 80% in the over 65 age group). It could be hypothesised that non-plantar lesions respond more readily as the epidermis in these areas is thinner, facilitating better penetration of microwave energy to the basal keratinocyte layer where HPV infection is most resilient, and where interaction with the cutaneous immune system is the most closely aligned thereby promoting a immune based curative process not reliant on necrosis by ablation or cryotherapy.

For patients with disorders affecting the immune system it is generally expected that cure rates may be lower. However, although we did find that clearance rates were lower, the clearance rate of 58.6% in those with autoimmune diseases and the immunocompromised (61.3%) was still favourable. Patients with diabetes may suffer prolonged wart infection ^7^ and may be precluded from traditional treatments such as topical acids. Heating of HPV lesions has been found to be highly effective at clearing genital warts in patients with diabetes ^8^. 2.4% of patients treated in this cohort were reported to have diabetes but this did not appear to affect the treatment outcome with 79.6% of patients reported to have wart clearance following microwave treatment, a rate equivalent to non-diabetic individuals.

Pain is an issue associated with most wart treatments and for some, microwave treatment can be uncomfortable. Most clinicians employed methods to mitigate treatment pain and approximately a third recommend over the counter analgesics such as paracetamol to their patients. The incidence of prolonged post-operative pain lasting over 4 weeks was found to be very low, reported by 4/93 respondents. Known side effects with microwave treatment include localised pain, prolonged pain, bruising or ulceration to the treated area and their occurrence increases with the energy dosage employed and the anatomical area treated.

There were no adverse events evident within the data which were deemed to be reportable to the regulatory authorities. but known side effects (ulceration, blister, haematoma) were reported and overall, reported patient satisfactions was high.

## Limitations

The data captured within this survey reflects questionnaire responses from practicing clinicians and is therefore subject to bias intrinsic to retrospective analyses and questionnaire data. To limit the influence of such bias on data interpretation, we assumed that the proportion of cleared warts would be more accurate than the absolute number. Therefore, for the analysis of plantar and common warts we report here the median reported clearance rates, rather than the cure rate calculated by the total number of lesions cleared per lesions treated (cohort clearance). However, the difference in result between either approach is minimal. For smaller datasets, e.g. immunosuppressed, autoimmune disease and diabetic patients, the numbers treated were fewer, thus the reported ‘cohort clearance’ is presented. Whilst every effort was made to gather a complete dataset only 58.5% of questionnaires were returned. In the questionnaire, responders were advised to input all case data and refer to medical notes, but it is not possible to validate the data returned to us, which could be subject to recall bias and reporting bias. However, we feel we have approached these data in a very stringent manner, reducing the impact of bias. Comparative data of responders versus non-responders to the questionnaire could reveal no significant differences between the two groups based on geography of the clinic, length of time the clinic has carried the treatment, socio-economic distribution or average usage of the device.

## Conclusion

This survey of microwave treatment for 8506 adult wart patients has shown that the treatment represents a valuable tool in the armamentarium for treatment of plantar and common warts. A randomized controlled trial would fully assess the efficacy of microwave treatment.

## Data Availability

All data produced in the present study are available upon reasonable request to the authors

