## Supplementary File 1 for "Post marketing surveillance for Microwave Treatment of Plantar and Common Warts in Adults"

**Copy of the Questionnaire**

Q1. General Information Address etc

Q2. How long have you been using Swift?

Q3. How many patients do you treat using Swift during an average week?

Q4. How happy are you with the treatment overall?

Q5. How happy have your patients been with the treatment overall?

Q6. Has Swift had a positive impact on your business?

Q7. How likely is it that you would recommend the Swift System to a friend or colleague?

Q8. Please briefly explain why you gave the Swift System this rating.

Q9. If a patient presents with a plantar wart and is a suitable candidate for Swift, what % of patients do you recommend the Swift treatment to?

Q10. How many patients will then choose the Swift treatment over other options?

Q11. For suitable patients who do not choose Swift, what are the cited reasons?

Q12. Efficacy in Adults - Under 65sHow many plantar wart patients have completed treatment to date? Please enter the number of adults (under 65):

Q13. How many plantar wart patients, that have completed treatment, have resolved? Please enter the number of adults (under 65):

Q14. How many common (non-plantar) wart patients have completed treatment to date? Please enter the number of adults (under 65):

Q15. How many common (non-plantar) wart patients, that have completed treatment, have resolved? Please enter the number of adults (under 65):

Q16. Adults Over 65How many plantar wart patients have completed treatment to date? Please enter the number of adults (over 65):

Q17. How many plantar wart patients, that have completed treatment, have resolved? Please enter the number of adults (over 65):

Q18. How many common (non-plantar) wart patients have completed treatment to date? Please enter the number of adults (over 65):

Q19. How many common (non-plantar) wart patients, that have completed treatment, have resolved? Please enter the number of adults (over 65):

Q20. Efficacy in Children How many plantar wart patients have completed treatment to date? Please enter the number of children (under 16):

Q21. How many plantar wart patients, that have completed treatment, have resolved? Please enter the number of children (under 16):

Q22. How many common (non-plantar) wart patients have completed treatment to date? Please enter the number of children (under 16):

Q23. How many common (non-plantar) wart patients, that have completed treatment, have resolved? Please enter the number of children (under 16):

Q24. Do you treat all lesions present, or primary lesions only, leaving satellite lesions without treatment?

Q25. If only primary lesions are treated, how many patients have seen satellite lesions resolve or improve without treatment?

Q26. OR provide a percentage - If only primary lesions are treated, how many patients have seen satellite lesions resolve or improve without treatment (%)?

Q27. How many treatments does a typical patient receive before resolution?

Q28. How many weeks do you leave between treatments?

Q29. How many months do you wait to review after the final treatment?

Q30. Do patients receive any other treatment (either clinically, or self-administered) while receiving Swift treatment?

Q31. If yes, do you see any change in efficacy?

Q32. Have you seen any instances of reoccurrence or reinfection (in the same area that was previously treated using Swift)?

Q33. If yes, how many patients have had instances of reoccurrence or reinfection (in the same area that was previously treated using Swift)?

Q34. What % was reoccurrence (rather than reinfection)?

Q35. Previous data collection showed clinicians typically using a protocol of 8-10W for 2 seconds, 3-5 repetitions, 4 weeks apart. Do you follow this protocol?

Q36. If no, please provide an alternative protocol for a typical, healthy patient (i.e. non-diabetic, over the age of 16, non-mosaic) Please enter the power used, in Watts:

Q37. If no, please provide an alternative protocol for a typical, healthy patient (i.e. non-diabetic, over the age of 16, non-mosaic) Please enter the time used, in seconds:

Q38. If no, please provide an alternative protocol for a typical, healthy patient (i.e. non-diabetic, over the age of 16, non-mosaic) Please enter the number of repetitions over the same spot:

Q39. If no, how many weeks do you leave between treatments?

Q40. If no, how many months do you wait to review after the final treatment?

Q41. How often do you use this protocol? (%)

Q42. In certain circumstances, would you use a different protocol?

Q43. If yes, please provide an alternative protocol - Please enter the power used, in Watts:

Q44. If yes, please provide an alternative protocol - Please enter the time used, in seconds:

Q45. If yes, please provide an alternative protocol - Please enter the number of repetitions over the same spot:

Q46. How many weeks do you leave between treatments?

Q47. How many months do you wait to review after the final treatment?

Q48. How often do you use this protocol? (%)

Q49. Under what circumstances would you use this protocol?

Q50. What % of Swift patients are (please enter a number):

Q51. Section 4.1: Immune-compromised patients Have you treated patients who are immune compromised?

Q52. How many immune compromised patients have completed treatment to date?

Q53. How many immune compromised patients have resolved?

Q54. Section 4.2: Children How old is the youngest patient you've treated?

Q55. How many treatments does a typical patient under the age of 16 receive before resolution?

Q56. In your experience, do children resolve quicker than adults?

Q57. Previous data collection showed clinicians typically using a protocol of 8-10W for 2 seconds, 3-5 repetitions, 4 weeks apart. Do you follow this protocol for a typical, healthy patient under the age of 16?

Q58. If no, please provide an alternative protocol -Please enter the power used, in Watts:

Q59. If no, please provide an alternative protocol -Please enter the time used, in seconds:

Q60. If no, please provide an alternative protocol -Please enter the number of repetitions over the same spot:

Q61. If no, how many weeks do you leave between treatments?

Q62. If no, how many months do you wait to review after the final treatment?

Q63. Section 4.3: Diabetic patients Have you treated any diabetic patients?

Q64. How many diabetic patients have completed treatment to date?

Q65. How many diabetic patients have resolved?

Q66. Section 4.4: Auto-immune patients Have you treated any auto-immune patients?

Q67. How many auto-immune patients have completed treatment to date?

Q68. How many auto-immune patients have resolved?

Q69. Do you treat any of these conditions?

Q70. Have you had any patients suffer prolonged pain (more than 4 weeks)?

Q71. If yes, how many patients have suffered prolonged pain?

Q72. Have you noticed any rare or serious side effects/adverse events? Please provide details below:

Q73. Did you notify Emblation of these events?

Q74. Have you used Swift on patients with metal implants?

Q75. Do you actively seek to manage pain?

Q76. If yes, how do you seek to manage pain?

Q77. Do you recommend painkillers post-treatment?

Q78. If yes, what do you recommend?

Q79. Is your management of pain during treatment different in children? Please provide details of pain management techniques used:
